# Population-based Oral Cancer Service Screening Disrupted by COVID-19 Pandemic: Observational and Simulation Study

**DOI:** 10.1101/2022.05.03.22274618

**Authors:** Chiu-Wen Su, Yen-Tze Liu, Amy Ming-Fang Yen, Han-Mo Chiu, Tony Hsiu-Hsi Chen, Tsui-Hsia Hsu, Ming-Yueh Shih, William Wang-Yu Su, Sam Li-Sheng Chen

## Abstract

**Background:** It is important for understanding the impact of COVID-19 pandemic on the missing opportunity for the early detection of oral cancer. This study aimed to assess the impact of COVID-19 pandemic on the existing population-based oral cancer (OC) service screening program in Taiwan.

**Methods:** Before and after COVID-19 pandemic design was used to assess the impact of COVID-19 on the reduction of screening rate, referral rate, and the effectiveness of this OC service screening. Data and analysis after pandemic covered non-VOC period in 2020 and VOC period in 2021 compared to the historical control before pandemic in 2019.

**Results:** The screening rate decreased substantially from 26.6% before COVID-19 in 2019 to 16.7% in 2020 and 15.3% in 2021 after pandemic. The reduction of screening rate varied with months, being the most remarkable decline in March (RR=0.61, 95% CI (0.60-0.62)) and June (RR=0.09, 95% CI (0.09-0.10)) in 2021 compared with January. The referral rate was stable at 81.5% in 2020 but it was reduced to 73.1% in 2021. The reduction of screening and referral rate led to the attenuation of effectiveness of advance cancer and mortality attenuated by 4% and 5%, respectively.

**Conclusion:** COVID-19 pandemic disrupted the screening and the referral rate and further led to statistically significant reduction in effectiveness for preventing advanced cancer and death. Appropriate prioritized strategies must be adopted to ameliorate malignant transformation and tumor upstaging due to deference from participation in the screening.

**Funding:** This study was financially supported by Health Promotion Administration of the Ministry of Health and Welfare of Taiwan (A1091116).

## INTRODUCTION

The emerging SARS-COV-2 has resulted in global public health crisis since mid-March, 2020 when the World Health Organization declared it as a pandemic infectious disease. As of August 22, 2021, the disease has spread over to 220 countries and regions with more than 212 million confirmed cases and 4.4 million deaths [1]. COVID-19 pandemic gives an unprecedented challenge for healthcare system in every country worldwide with many non-essential clinical visits and admissions that have been deferred [2]. The COVID-19 pandemic is still an ongoing transmission under the threaten of variant of concern (VOC) and interest in many countries. Although available anti-virus therapy or vaccination is provided, containment measures including the quarantine and the isolation of suspected individuals, social distancing, and personal and environmental sanitation are still widely adopted in many countries. In clinical settings, additional containment measures were applied to reducing the wide spread of transmission. Taking an example of dental practice, except emergencies, other dental activities have been suspended in most of affected countries [3, 4, and 5]. During the COVID-19 pandemic, some important health issues, like early detection of oral cancer may be neglected by the public attention. As screening with oral visual inspection for high-risk subjects is an effective means for oral cancer prevention worldwide [6, 7] that has shown the reduction of 21% advanced stage and 26% mortality of oral cancer. Thus, population-based oral cancer screening program had been considered as an important secondary prevention strategy due to the high disease burden of oral cancer. Although early detection of oral potentially malignant/malignant lesions contributes to reducing mortality and advanced cancers, non-participants in the screening [6] or delay in treatment [8] are associated with unfavorable outcome [9, 10]. It is of great interest to investigate the impact of COVID-19 pandemic on the missing opportunity for the early detection of oral cancer.

Although Taiwan has several community-acquired outbreaks with relatively lower confirmed cases and low case-fatality rates due to a fast response to COVID-19, routine health care service has been still deferred even when the service has remained fully functioning and accessible [11]. One of possible services that may be affected by COVID-19 pandemic [12] is pertaining to population-based service screening program whereby a proportion of residents would be invited to attend the screening on population level. Under the well-controlled circumstance of COVID-19 in Taiwan, there is generally no barrier for participating in the screening when invited, being referred for confirmatory diagnosis, or receiving subsequent treatment, whether basic screening characteristics such as screening rate and referral rate have been affected is worthy of being investigated.

Both lower screening rate and referral rate may further result in advanced stage of oral cancer and lead to the elevated mortality. In this study, we reported how these basic screening characteristics (such as screening rate and referral rate) and effectiveness of the population-based oral cancer screening program would be disrupted by COVID-19 pandemic.

## MATERIALS AND METHODS

### Population-based screening program for oral neoplasm

In Taiwan, population-based screening for oral neoplasm with oral visual inspection has been offered for subjects with habits of cigarette smoking and/or betel quid chewing since 2004. The details of this population-based screening program have been fully described elsewhere [6]. In brief, the cigarette smoker or betel quid chewer aged 30 years or over were invited to attend a biennial screening program with oral visual inspection by well-trained dentists or physicians to carry out on the ground of early detection of oral pre-malignancy and oral cancer. After screening, the participants with oral potentially malignant disorders or suspected malignancy were referred to the specialists in the hospitals for confirmatory pathological examination. The study approved by the Research Ethics Committee of National Taiwan University Hospital prior to data collection pursuant to the regulation of the Institutional Review Board (202103098W). In this study, the change of screening rate and referral rate were used to measure the impact of COVID-19 on oral cancer screening.

### Study Design

Based on this population-based screening data over the past one year, we found January and March had the most participants between 70,000 to 90,000 in 2019. The monthly participants less than 70,000 in February (Chinese new year festival) and each month after April in 2019 (Table 1). January had the most referrals around 7,000 but December had the less referrals around 2000 in 2019. For other months around 3,000 to 5,000 positive cases each month were referred to have confirmatory diagnosis (Table 2). The monthly referral rates were steady in 2019. Based on these screening characteristics, we used a monthly series of before and after quasi-experimental design to compare both screening rate and referral rate in 2020 (non-VOC period) and from January to June in 2021 (VOC period) with those of the historical control of the contemporaneous period in 2019 (non-COVID-19 period).

**Table 1.** Number of participants and relative rates regarding the change of the screening rate before (2019) and after (2020 and 2021) COVID-19 pandemic in oral cancer screening program in Taiwan.

| Month | Year |  |  |  |  |  |  |  |  |  |
| --- | --- | --- | --- | --- | --- | --- | --- | --- | --- | --- |
|  | 2019 |  | 2020 |  |  |  | 2021 |  |  |  |
|  | Number of participants | Screening rate | Number of participants | Screening rate | RR | (95%CI) | Number of participants | Screening rate | RR | (95%CI) |
| **January** | 90,914 | 6.2% | 71,366 | 4.8% | 1.00 |  | 59,226 | 4.0% | 1.00 |  |
| **February** | 56,297 | 3.9% | 46,416 | 3.1% | 1.05 | (1.04-1.06) | 39,272 | 2.6% | 1.07 | (1.06-1.09) |
| **March** | 70,687 | 4.8% | 33,812 | 2.3% | 0.61 | (0.60-0.62) | 55,240 | 3.7% | 1.20 | (1.19-1.21) |
| **April** | 61,405 | 4.2% | 27,023 | 1.8% | 0.56 | (0.55-0.57) | 46,365 | 3.1% | 1.16 | (1.15-1.17) |
| **May** | 59,185 | 4.0% | 30,679 | 2.1% | 0.66 | (0.65-0.67) | 24,978 | 1.7% | 0.65 | (0.64-0.66) |
| **June** | 50,882 | 3.5% | 37,794 | 2.6% | 0.95 | (0.94-0.96) | 3,051 | 0.2% | 0.09 | (0.09-0.10) |
| **July** | 46,537 | 3.2% | 43,401 | 2.9% | 1.19 | (1.18-1.21) | - | - | - |  |
| **August** | 42,115 | 2.9% | 41,450 | 2.8% | 1.26 | (1.24-1.27) | - | - | - |  |
| **September** | 35,195 | 2.4% | 37,807 | 2.6% | 1.37 | (1.35-1.39) | - | - | - |  |
| **October** | 36,165 | 2.5% | 34,718 | 2.3% | 1.22 | (1.21-1.24) | - | - | - |  |
| **November** | 32,128 | 2.2% | 29,711 | 2.0% | 1.18 | (1.16-1.2) | - | - | - |  |
| **December** | 22,007 | 1.5% | 19,825 | 1.3% | 1.15 | (1.13-1.17) | - | - | - |  |
| **Jan. to June** | 389,370 | 26.6% | 247,090 | 16.7% |  |  | 228,132 | 15.3% |  |  |
| **Jan. to Dec.** | 603,517 | 41.2% | 454,002 | 30.6% |  |  | - | - |  |  |

**Table 2.** Referral rate and relative rates regarding the change of the screening rate before (2019) and after (2020 and 2021) COVID-19 pandemic in oral cancer screening program in Taiwan.

| Month | Year |  |  |  |  |  |  |  |  |  |  |  |  |
| --- | --- | --- | --- | --- | --- | --- | --- | --- | --- | --- | --- | --- | --- |
|  | 2019 |  |  | 2020 |  |  |  |  | 2021 |  |  |  |  |
|  | N | n | Referral<br>rate | N | n | Referral<br>rate | RR | (95%CI) | N | n | Referral<br>rate | RR | (95%CI) |
| January | 6,849 | 5,573 | 81.4% | 4,853 | 3,883 | 80.0% | 1.00 |  | 4,756 | 3,585 | 75.4% | 1.00 |  |
| February | 4,411 | 3,511 | 79.6% | 3,416 | 2,834 | 83.0% | 1.06 | (1.01-1.11) | 3,225 | 2,373 | 73.6% | 1.00 | (0.94-1.05) |
| March | 5,628 | 4,509 | 80.1% | 2,683 | 2,213 | 82.5% | 0.99 | (0.94-1.04) | 5,085 | 3,587 | 70.5% | 0.95 | (0.90-1.00) |
| April | 5,178 | 4,144 | 80.0% | 2,158 | 1,801 | 83.5% | 1.01 | (0.96-1.07) | 4,286 |  |  |  |  |
| May | 5,034 | 4,056 | 80.6% | 2,725 | 2,291 | 84.1% | 1.00 | (0.95-1.05) | 2,233 |  |  |  |  |
| June | 4,459 | 3,541 | 79.4% | 3,168 | 2,555 | 80.7% | 0.97 | (0.93-1.02) | 392 |  |  |  |  |
| July | 4,003 | 3,101 | 77.5% | 3,753 | 3,065 | 81.7% | 1.04 | (0.99-1.09) | - |  |  |  |  |
| August | 3,549 | 2,743 | 77.3% | 3,452 | 2,735 | 79.2% | 0.97 | (0.92-1.03) | - |  |  |  |  |
| September | 2,969 | 2,252 | 75.9% | 3,114 | 2,456 | 78.9% | 1.01 | (0.96-1.07) | - |  |  |  |  |
| October | 3,020 | 2,459 | 81.4% | 2,595 | 2,075 | 80.0% | 0.94 | (0.89-1.00) | - |  |  |  |  |
| November | 2,661 | 2,143 | 80.5% | 2,634 | 1,999 | 75.9% | 0.96 | (0.90-1.02) | - |  |  |  |  |
| December | 2,064 | 1,689 | 81.8% | 2,111 | 1,589 | 75.3% | 0.98 | (0.91-1.05) | - |  |  |  |  |
| Jan. to March | 16,888 | 13,593 | 80.5% | 10,952 | 8,930 | 81.5% |  |  | 13,066 | 9,545 | 73.1% |  |  |
| Jan. to Dec. | 49,825 | 39,721 | 79.7% | 36,662 | 29,496 | 80.5% |  |  | - | - | - |  |  |

n: number of positive cases who were compliant to referral
N: number of subjects with positive finding after screening

As we do not have the control group as seen in population-based randomized controlled trial, it is very difficult to estimate the risks of advanced oral cancer or deaths in the absence of screening. A multi-state Markov natural history model as an alternative approach was applied to simulating the cancer risks for subjects with and without screening.

### Outcome Measurement and Data Collection

#### Indicators for Screening Program

The two major in relation to the outcomes of screening process used in this study were screening rate and referral rate. Screening rate was defined as the rate of number of participants to population aged over 30 years with cigarette smoking and/or betel nut chewing. The referral rate was defined as number of positive cases with confirmatory diagnosis.

#### Data Collection of Oral Cancer Screening

Those who had the clinical diagnosis with suspicious oral malignancy were referred to confirm the diagnosis or oral cancer by tissue biopsy. Such a clinical diagnosis may include a persistent white or red lesion, a non-hearing ulcer for more than two weeks, an irregular surface lesion inside the oral cavity, or any unexplained lymph adenopathy during the oral visual inspection was defined as positive case. Needle or incisional biopsy would be needed before a confirmed diagnosis. Individual screen data from clinics or hospitals are timely uploaded and transferred to a centralized database via web-based information system to facilitate the process and outcome evaluation of screening. Data on number of targeted screen populations, number of participants, and number of referrals were collected from Taiwanese Oral Cancer Screening Database.

#### Data Collection of COVID-19

COVID-19 reported data used were obtained from the Taiwan National Infectious Disease Statistics System (TNIDSS) [13], which was maintained by the Taiwan Centers for Disease Control. They daily reported confirmed cases were collected.

### Statistical Analysis

The chi-square test was used to assess the differences in both screening rate and referral rate before and after COVID-19. The statistically significant level was set as 0.05. For the calculation of relative rate for screening rate change, the monthly screening rate in 2020 was compared with that in 2019. The relative ratio of screening rate change in the epidemic period was reckoned on the basis of the relative rate of screening from pre-epidemic period in January. The 95% confidence intervals of relative rate were calculated.

We simulated a cohort based on a three-states natural history model (from normal, oral potentially malignant disorder (OPMD) state to cancer state) incorporating oral behaviors [14] to project the effectiveness of oral cancer screening on advanced cancer incidence and oral cancer mortality. The model assumes the cancer would be developed following the pathway of malignant transformation through OPMD. The disease progression from OPMD to cancer depends on the use of oral behaviors including alcohol, tobacco, and betel nut [6]. In addition, some with these oral habits are associated with malignant transformation [14] to oral cancer. Moreover, some cases of oral cancer may even progress to more advanced stages. It should be noted that the survival rate after diagnosis depends on the tumour stage. The early detection by screening results is to reach the goal of stage shifting toward stage 0 or 1[6, 7].

We firstly used a hypothetical cohort without any intervention to simulate and project the expected number of advanced oral cancer and then changed the screening rate from pre-pandemic period to pandemic period during 6-year follow-up to estimate oral cancer deaths. The parameters used in the current simulation are shown in Supplementary Table 1. The statistical software used for the analysis was SAS 9.4.

## RESULTS

### The impact of delayed screening on basic screening characteristics

Figure 1 shows the epidemiological profile of confirmed cases of COVID-19 from January, 2020 to June, 2021 in Taiwan with a peak on March 16^th^, 2020 resulting from imported cases and another peak on May 20^th^, 2021 resulting from regional outbreak. After a series of containment measures such as social distancing, there was a substantial decline in confirmed cases of COVID-19. Since our nationwide screening program has started since 2004 the monthly number of eligible residents invited to screen during early half of the year has a consistent pattern based on the data of 2019 prior to COVID-19 pandemic as shown in Table 1. The figure had the highest in January, dropped to the lowest in February, then rose in March, and slightly fell after April. For ease of presentation, Figure 2 (A) shows the comparison of number of screenee between 2019 and 2021. The trends decreased substantially after 2019, starting with a screening rate of 6.2% in Jan 2019, 4.8% in 2020, 4.0% in 2021 (Figure 2(B)). The referral rate also decreased to 80.0 % in January 2020 and 75.4% in January 2021 compared with the corresponding figure of 81.4% in 2019) (Figure 2(C)).

**Figure 1.**
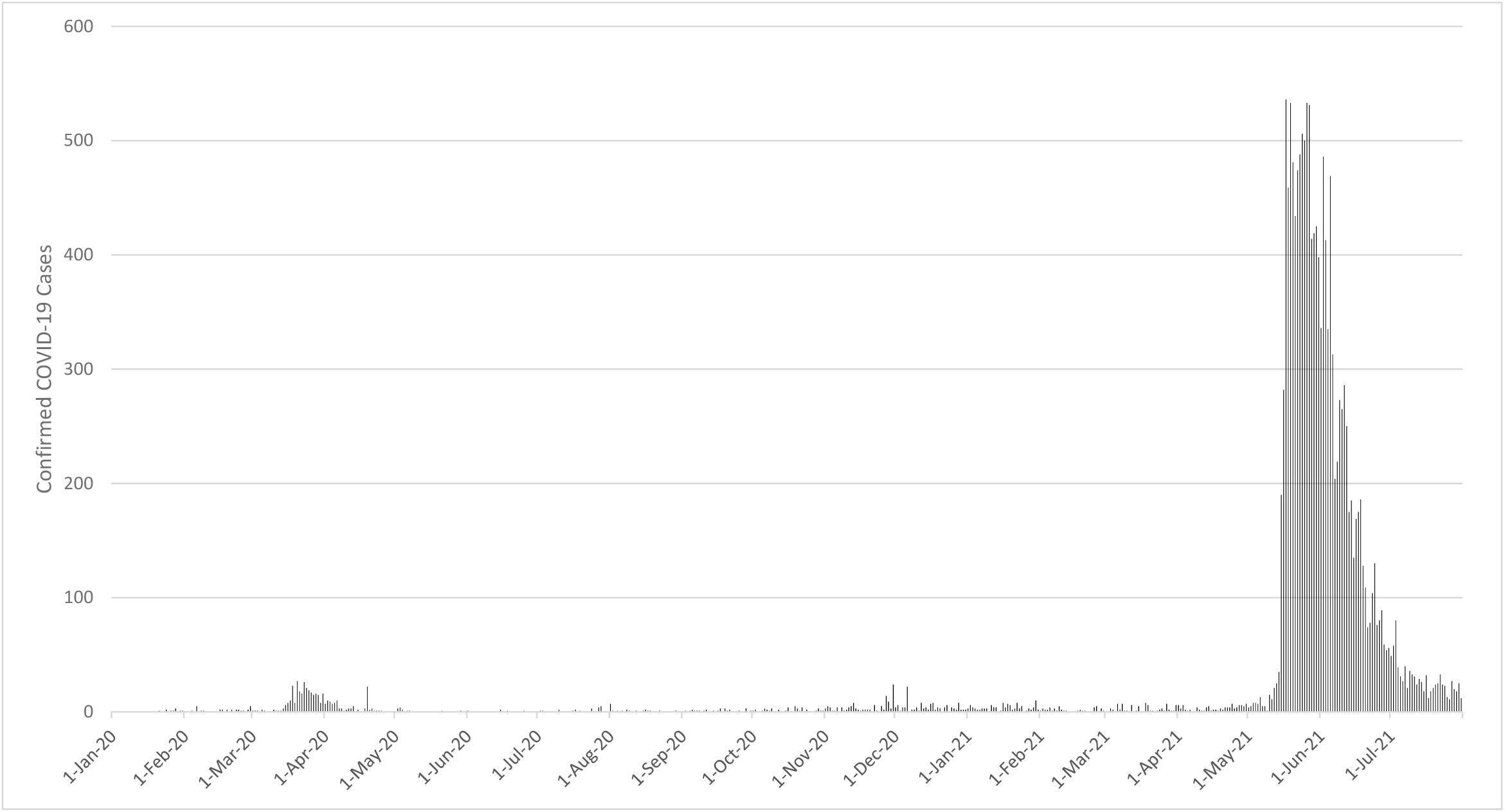
Number of confirmed COVID-19 cases in Taiwan during the COVID-19 pandemic.

**Figure 2.**
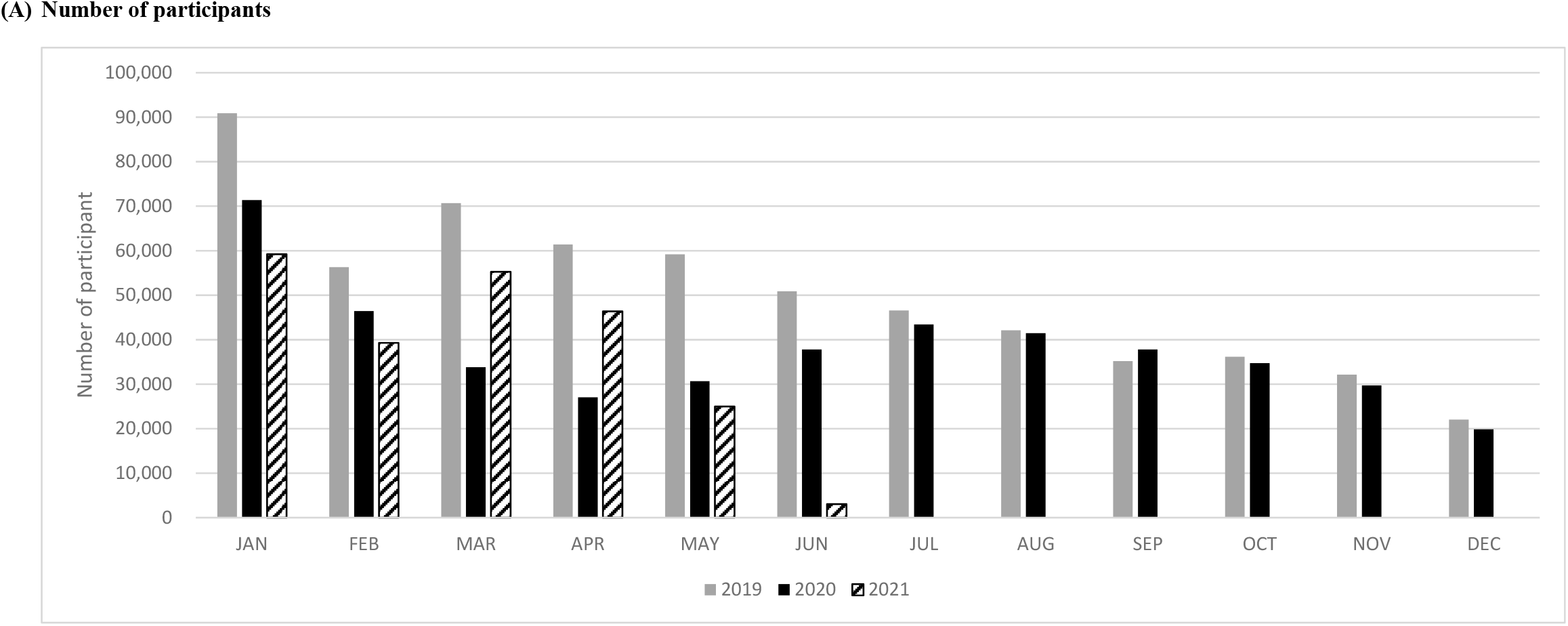
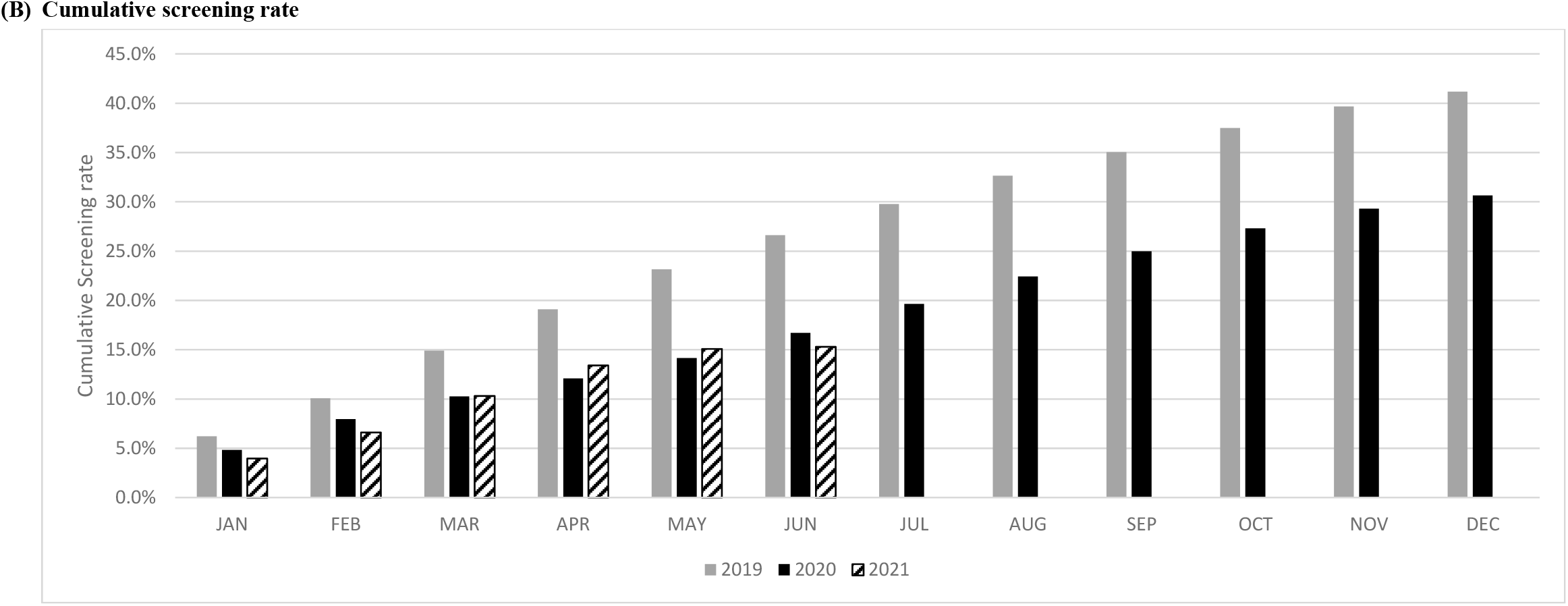
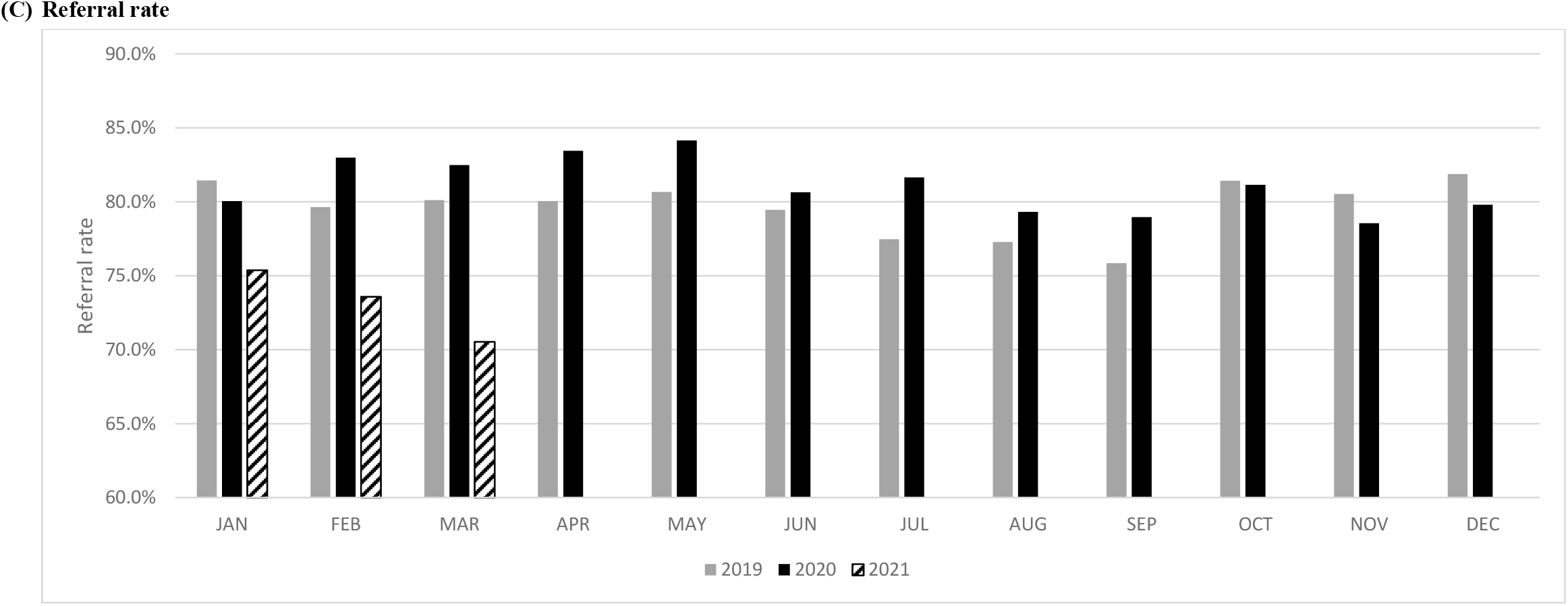
Number of participants, screening rate and referral rates before (2019) and after (2020 and 2021) COVID-19 pandemic in oral cancer screening program in Taiwan.

Table 1 shows the results of relative rates regarding the comparison of the monthly screening rate from January to December between 2019 and 2020 and from January to June in 2021 taking the figure of January as the baseline group. There was lacking of reduction in February whereas the reduction of the screening rate in 2020 versus 2019 was increased from 39% (RR=0.61, 95% CI (0.60-0.62)) in March, 44% in April (RR: 0.56, 95% CI (0.55-0.57)), and to 34% in May (RR: 0.66, 95% CI (0.65-0.67)), (Table 1). The screening rate increased from July onwards. The referral rate did not change for each month. (Table 2). However, the reduction of the screening rate in 2021 versus 2019 was dramatically increased from 35% (RR=0.65, 95% CI (0.64-0.66)) in May and 91% in June (RR: 0.09, 95% CI (0.09-0.1)). Compared with the 81.5% of referral rate between January and March before 2021, the average referral rate decreased to 73.1% in the corresponding period of 2021.

### Simulation of the impact of oral cancer screening delays on cancer detection and related death

Given 26.6% of screening rate and 80.5% of referral rate in the period between January and June in 2019, oral cancer screening reduced advanced cancer and mortality by 29% (24%-33%) and 17% (11%-23%), respectively, over 6 years, whereas the decline in screening rate (16.7%) and referral rate (81.5%) in 2020 due COVID-19 pandemic may compromise the effectiveness by 4%, resulting in 25% (20%-29%) in advance cancer reduction and 13% (7%-19%) in mortality reduction (Table 3). The similar result shows that the delayed screening and referrals may compromise the effectiveness by 5% in 2021.

**Table 3.**
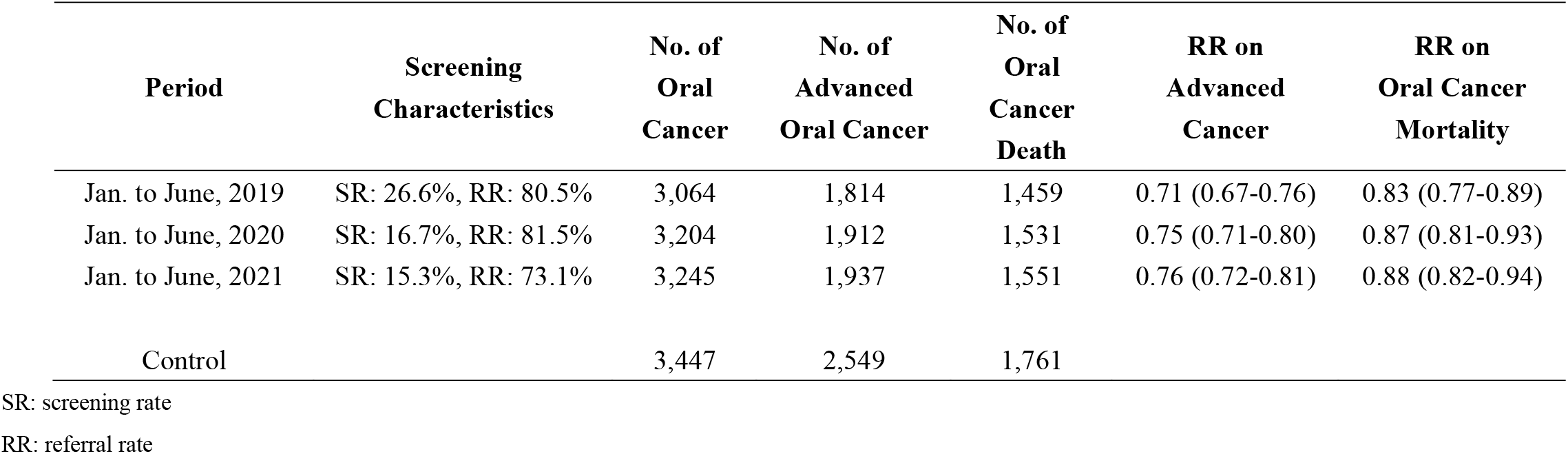
Simulated effectiveness of oral cancer screening at reducing advanced oral cancer and oral cancer mortality during the COVID-19 pandemic.

| Period | Screening Characteristics | No. of Oral Cancer | No. of Advanced Oral Cancer | No. of Oral Cancer Death | RR on Advanced Cancer | RR on Oral Cancer Mortality |
| --- | --- | --- | --- | --- | --- | --- |
| Jan. to June, 2019 | SR: 26.6%, RR: 80.5% | 3,064 | 1,814 | 1,459 | 0.71 (0.67-0.76) | 0.83 (0.77-0.89) |
| Jan. to June, 2020 | SR: 16.7%, RR: 81.5% | 3,204 | 1,912 | 1,531 | 0.75 (0.71-0.80) | 0.87 (0.81-0.93) |
| Jan. to June, 2021 | SR: 15.3%, RR: 73.1% | 3,245 | 1,937 | 1,551 | 0.76 (0.72-0.81) | 0.88 (0.82-0.94) |
| Control |  | 3,447 | 2,549 | 1,761 |  |  |

SR: screening rate
RR: referral rate

## DISCUSSION

Our data show significant 24.8% reduction in number of participating (603,517 in 2019 and 454,002 in 2020) subjects and 41.4% reduction in number of participating (389,370 in first half of 2019 and 228,132 in first half of 2021) subjects and 8% difference in the referral rate in 2021 for population-based oral screening in Taiwan during the COVID-19 pandemic. Since there is no obvious mitigation of spread worldwide, a continual decrease in these indexes can be foreseen. A retrospective study from Germany also found cancer diagnosis delay with the high proportion of the high tumor stages during the lockdown in COVID-19 pandemic but there was no higher incidence of oral cancer due to the shorter observed time [4]. The subjects with oral potentially malignant lesions may yield a 3.5% (range 0.3-34%) risk of annual malignant transformation rate [15] while those with occult oral malignancy may have caner progression or up-staging if not detected. Overall, the high-risk subjects who do not participate in the screening have a 1.26-fold relative risk of been diagnosed at advanced stage and a 1.35-fold of mortality from oral cancer [6]. Consequently, we could expect a higher incidence or more advanced oral cancer as shown in our simulated results if the screening program is not able to proceed as usual. Based on the current scenario in our screening program, the reduction of 4∼5% in both advanced cancer and mortality would be expected. A recent modelling study also revealed a 6% lower effectiveness in reducing colorectal cancer mortality by prolonged pauses of colorectal cancer screening [16].

With the unprecedented challenge from the COVID-19, the US Centers for Disease Control has recommended to cease elective care and provide only urgent visits and emergent procedures [17]. However, the healthcare capacity in Taiwan was never strained owing to proactive border control, active surveillance, rapid quarantine, and resource allocation to prevent the spread of COVID-19 in communities, which was executed by the Central Epidemic Command Center after the first confirmed case on January 21, 2020 [18]. With insufficient COVID-19 vaccine coverage, all people were asked to report travel, occupation, contact, and cluster histories, wear mask, sanitize hands, and have body temperature checked at the quarantine stations of every medical facilities [19]. As such, the long queue, waiting, and crowding may hinder subjects from participating in the screening. Also, the information of COVID-19 pandemic is incessantly emphasized and widespread through media and social networks, which further reduces the willingness to visit healthcare facilities owing to the fear of been transmitted. Some may even deem physicians as a source of contagion and avoid been referred after positive screening results [17].

The high viral load in upper aerodigestive tract [20] and the proximity during screening render the physicians to infection [21]. The use of tongue depressor to facilitate the inspection sometimes induced cough which created aerosols and caused increased risk of transmission [22]. As the first reported physician fatality related to COVID-19 in Wuhan, China was an otolaryngologist [17], all these concerns further diminish the physicians’ willingness to perform screening. Although there is no guideline regarding oral cancer screening during the pandemic, societies do recommend “no routine head and neck examination” [20] and postpone all non-essential dental exams and procedures [3]. However, the enormous susceptive cases after screening waiting for referral reflected that the screening should not be ignored during the pandemic [2]. The tele-consultation or self-examination by high-end mobile technology [23] might be alternative solutions when we are faced with new viral variants or new emerging infectious diseases.

To date, it is unclear whether the COVID-19 would ease or becomes long-lasting threats to human, we should not let the pandemic disrupt the screening. Approaches to ensure safety for the physicians and the subjects are essential, such as via providing adequate personal protective equipment or rapid throat swab tests [24]. Last, it is crucial to assure the public the importance to resume participating in oral cancer screening and complying with referral if the result is positive.

## CONCLUSION

In conclusion, both of the screening rate and referral rate were significantly reduced during the COVID-19 pandemic. High effectiveness of population-based oral cancer screening is not achieved if both cannot be recovered. The screening organizer should continuously coordinate with the screening institutes to setup the priority for screening and apply modern technologies to overcome potential barriers in the screening.

## Data Availability

Data and program are available at online, link: https://datadryad.org/stash/share/KMt0vv6jpHhdJlozlY4WkEOva17nwTw7ePP-tDcMtjE

## Acknowledgement

This research was funded by the Health Promotion Administration, Ministry of Health and Welfare of Taiwan under the program of “Breast cancer, oral cancer, and colorectal cancer screening data monitoring and evaluation research program, Year 2021” (A1091116).

